# Clinical presentation of severe malaria in children who received the RTS,S/AS01_E_ malaria vaccine, seasonal malaria chemoprevention or the combination of both interventions in Burkina Faso and Mali

**DOI:** 10.64898/2026.02.11.26344823

**Authors:** Djibrilla Issiaka, Issaka Zongo, Youssoufa Sidibe, Yves Daniel Compaoré, Rakiswendé Serge Yerbanga, Mahamadou Kaya, Charles Zoungrana, Romaric Oscar Zerbo, Oumar M Dicko, Youssouf Kone, Amadou Tapily, Seydou Traore, Koualy Sanogo, Modibo Diarra, Amadou Barry, Almahamoudou Mahamar, Alassane Haro, Ismail Thera, M Sanni Ali, Paul Snell, Jane Grant, Irene Kuepfer, Cynthia K. Lee, Christian F. Ockenhouse, Opokua Ofori-Anyinam, Paul Milligan, Issaka Sagara, Halidou Tinto, Jean Bosco Ouedraogo, Daniel Chandramohan, Brian Greenwood, Alassane Dicko

## Abstract

**Background:** Severe *Plasmodium falciparum* malaria is a leading cause of death in sub-Saharan Africa, with most deaths occurring in children younger than five years of age. The RTS,S/AS01_E_ (RTS,S) malaria vaccine delivered seasonally with Seasonal Malaria Chemoprevention (SMC) led to a two-third reduction in severe malaria and malaria deaths compared with either intervention given alone. The aim of this study was to assess the seasonal distribution and clinical presentation of children admitted in hospital with severe malaria who received RTS,S with or without SMC, and whether seasonal vaccination with RTS,S affected their distribution and clinical presentation.

**Methods:** We conducted a secondary ad-hoc analysis of hospital admissions in children aged 5 to 17 months of age when enrolled in the RTS,S + SMC trial in April 2017 in Houndé, Burkina Faso, and Bougouni, Mali, and who were followed until they reached five years of age.

**Results:** Three hundred and thirtythirty-seven serious adverse events were reported during the trial with 222 (65.9%) in Burkina Faso and 115 (34.1%) in Mali, and 48.1% (162/337) were due to severe malaria. The mean age of children with severe malaria was 2.9 years; 79.0% (128/162) of the cases occurred in children aged 3 years or less and 21.0% (34/162) in those aged 4 to 5 years. The most common presentation was severe anemia reported in 50% (81/162) of children, followed by repeated convulsions 25.3%, (41/162), prostration 11.1%, (18/162), hyperparasitaemia 8.0%, (13/162) and cerebral malaria 6.2%, (10/162). Severe malaria anemia was a more frequent form of severe malaria in children in the SMC alone arm 62.1%, (36/58) and lower in children in the SMC+ RTS,S arm 37.1%, (13/35), p=0.048. There was no significant difference in frequency of other clinical presentations between the study arms. Most severe malaria cases, 85.8% (139/162) occurred during the transmission season (July to December).

**Conclusions:** Severe anaemia was the most common presentation of severe malaria and was most frequent in the SMC alone arm. Children aged 3 years or less were the most affected and almost all the cases occurred between July and December.

**Trial registration:** ClinicalTrials.gov, NCT04319380

## BACKGROUND

In sub-Saharan Africa, falciparum malaria is a major cause of morbidity and mortality. Children under the age of five years account for the majority of cases of severe malaria and malaria deaths ^1^. Over the last two decades, malaria prevention in children has relied mainly on the use of insecticide-treated bed nets (ITNs) and seasonal malaria chemoprevention (SMC). The introduction of SMC, recommended by WHO in 2012 ^2^, has led to a substantial reduction in malaria infection and disease including severe malaria ^3,4^. This strategy is now widely implemented with about 228 million doses of SMC distributed to 54 million of children in 2024 according to a recent WHO report ^1^. Despite the wide implementation of SMC and continued improvement in access and coverage of ITNs, the burden of the malaria continues to be high in the areas of sub-Saharan Africa where malaria transmission is highly seasonal and there is a clear need for additional tools such as vaccines to reduce this burden.

RTS,S/AS01_E_ (RTS,S) was the first malaria vaccine to be recommended for widespread deployment by WHO in 2021^5^. A study conducted in Burkina Faso and Mali showed that RTS,S given as seasonal vaccination was not inferior to SMC and that the combination of the two interventions reduced hospital admissions and deaths due to severe malaria by two-thirds ^6,7^. The rationale for seasonal vaccination is that the administration of a malaria vaccine, such as RTS,S which provides only a limited period of high efficacy, at the beginning of the high-transmission season^8^ in areas where malaria incidence is highly seasonal, will provide maximum protection for children when they are at greatest risk of infection and severe disease.

The burden of severe malaria is influenced by malaria transmission intensity, seasonality and the use of preventive measures, all of which can influence malaria immunity, age distribution and the clinical presentation of the disease. In areas of high and perennial transmission, young children are the most vulnerable group whilst within areas of low seasonal transmission, immunity develops more slowly and the age distribution of severe malaria cases is broader ^9,10^ with more older children being affected. It has been reported in a systematic review of severe malaria in different settings in sub-Sharan Africa ^11^ that the median age for cerebral malaria cases was around 34 months in high-transmission settings; in low-transmission settings, the median age was nearly 49 months. This epidemiological shift was also reported in Eastern Uganda and other parts of Africa where the use of malaria control measures have altered transmission patterns, leading to increased severe cases among older children and adolescents ^12^.

In seasonal transmission settings, severe malaria anaemia (SMA) has been reported to be the most common clinical presentation in African children aged under 2 years, with cerebral malaria occurring later in children aged 3 to 4 years, ^13–16^ and that most of the cases occurred during the transmission season ^15,17^.The study described in this paper aimed to assess the seasonal distribution and clinical presentation of severe malaria admissions in children who received RTS,S with or without SMC in Burkina Faso and Mali, and whether seasonal vaccination with RTS,S affected their clinical presentation.

## METHODS

### Study sites

The study was conducted in the Houndé health district in Burkina Faso and in Bougouni and its surrounding villages in Mali. A description of the study sites was presented in a previous paper ^6^. Malaria transmission is highly seasonal, occurring from July to November, in both districts. The incidence of uncomplicated malaria in children under 5 years of age using an ITN and receiving SMC was estimated at 877.4 cases per 1,000 person-years in Burkina Faso and 865.2 cases per 1,000 person-years in 2016 in Mali. The incidence of malaria-related hospital admissions or deaths was 13.8 per 1,000 person-years in Burkina Faso, compared to 11.2 per 1,000 person-years in Mali ^15^.

### Overall study design

This study is a secondary analysis of data from the RTS,S-SMC trial (ClinicalTrials.gov, NCT04319380) already described by Dicko et al., 2024 ^6^. Briefly, after obtaining ethical and regulatory approvals and community consent for the trial, a census of all households within the study area in each country was undertaken in April 2017 to identify children aged 5 to 17 months of age. Eligible children were enrolled after obtaining individual informed consent from a parent or guardian. Children were randomized into one of three arms to receive: i) the RTS,S vaccine with placebo SMC, ii) SMC plus a control vaccine, or iii) RTS,S and SMC. The randomisation list was available to the study pharmacist but site investigators, including clinical staff, the laboratory staff, and the study support staff, were kept blinded to the group to which a child had been allocated. At the time of eligibility assessment, each study participant received an identification card with the child’s photograph and a reference code bar. The latter was used for each study procedure, laboratory blood collection, vaccination, or SMC administration.

### Interventions

Children in the RTS,S alone or RTS,S +SMC combined group received three priming doses of the RTS,S vaccine from April to June 2017 at monthly intervals followed by two booster doses in June 2018 and June 2019 prior to each malaria transmission season (initial period of trial). In 2020, the protocol was amended to allow the administration of the two interventions, singly or in combination, for a further two years (the extension period, ClinicalTrials.gov, NCT04319380.) and children in the RTS,S group alone or the RTS,S+SMC combined group received further booster doses of RTS,S in June 2020 and June 2021 (Figure 1). The children in the SMC alone group received three injections of rabies vaccine (Rabipur ^@^) from April 2017 to June 2017 and one injection of rabies vaccine in June 2018 and June 2019 prior to the malaria transmission season. In the extension period, the participants in the SMC alone group received tetanus or tetanus-diphtheria vaccine in June 2020 and June 2021. The SMC alone and the combined group received SMC with sulphadoxine-pyrimethamine and amodiaquine (SPAQ) with a monthly cycle each year. Four cycles of SMC were given according to the national guidelines of Mali and Burkina Faso from July to October, with an additional cycle being given in Burkina Faso in November of the last year of the extension study in line with national policy. All the children received an insecticide-treated net prior to the administration of the first dose of the vaccine in April 2017 and in June 2020.

**Figure 1:**
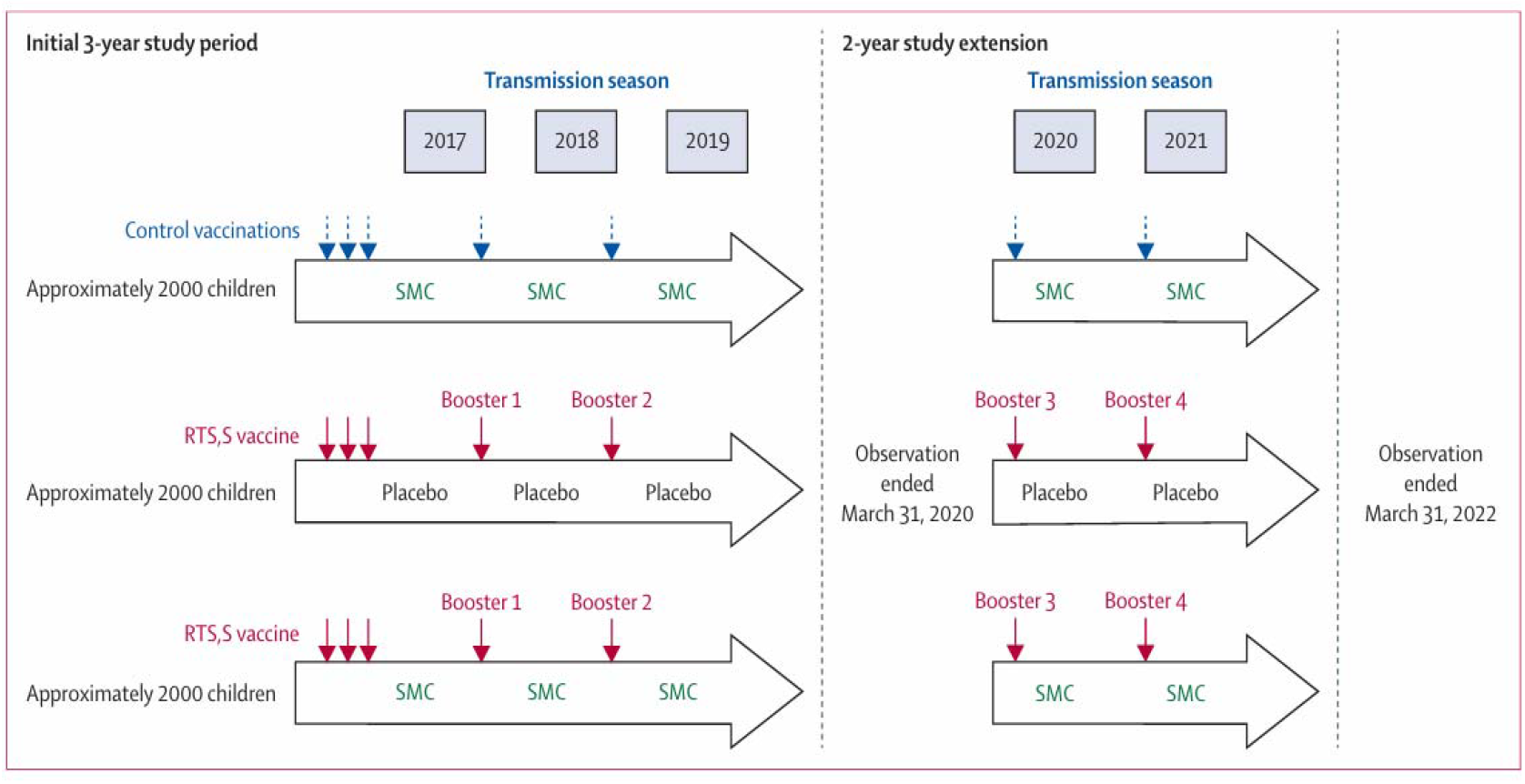
Study design (source: Dicko et al Lancet ID 2023).

### Surveillance for malaria

Children who attended a health facility with suspected malaria or a measured temperature of 37.5 degrees Celsius or higher, or a history of fever within the past 48 hours, were tested for malaria using a rapid diagnostic test (PfHRP2) and a blood film was collected for subsequent microscopic examination^18^. Serious adverse events (SAEs) were recorded throughout the trial.

All study children admitted to hospital were identified by the clinical staff based in these hospitals. Information on their clinical presentations was collected for each case of severe malaria. These clinical features were recorded in the SAE form based on the information provided by the parent since the start of the illness and on physical examination. Severe malaria cases considered for this analysis were defined using the WHO criteria ^19^ (Table1).

**Table 1.** Outline classification of severe malaria in children^19^.

|  |
| --- |
| Group 1 |
| Children at immediately increased risk of dying who require parenteral antimalarial drugs and supportive therapy |
| (a) Prostrated children (prostration is the inability to sit upright in a child normally able to do so, or to drink in the case of children too young to sit) |
| Three subgroups of increasing severity should be distinguished: |
| (i) Prostrate but fully conscious |
| (ii) Prostrate with impaired consciousness but not in deep coma |
| (iii) Coma (the inability to localize a painful stimulus) |
| (b) Respiratory distress (acidotic breathing): |
| (i) Mild — sustained nasal flaring and/or mild intercostal indrawing (recession) |
| (ii) Severe — the presence of either marked indrawing (recession) of the bony structure of the lower chest wall or deep (acidotic) breathing |
| Group 2 |
| Children who, though able to be treated with oral antimalarial drugs, require supervised management because of the risk of clinical deterioration but who show none of the features of Group 1 |
| (a) Children with a haemoglobin level < 5 g/dL or a haematocrit < 15% |
| (b) Children with 2 or more convulsions within a 24 h period |
| Group 3 |
| Children who require parenteral treatment because of persistent vomiting but who lack any specific clinical or laboratory features of Groups 1 or 2 |

Causes of deaths, whether in the hospital or outside of hospital were assessed by two clinicians. In the case of discrepancy, a third opinion was obtained from another clinician to reach a consensus diagnosis. Deaths that occurred outside health centers, investigated by verbal autopsy method in accordance with WHO guidelines ^20^, were not included in this analysis as biological confirmation could not be made.

### Data collection and statistical analysis

Data were collected on electronic case report forms and analysed using STATA version 15 (https://www.stata.com/stata.com/). Proportions were compared using Pearson Chi square or Fisher exact test for the comparison of proportions and Student t test was used for comparison of mean age between the two countries. The significance level was set at 5%.

## RESULTS

Three hundred and thirty-seven serious adverse events (SAEs), were reported during the five years of the trial, with 222 in Burkina Faso and 115 in Mali; 48.1% (162/337) were due to severe malaria; 69/162 (42.6%) in the RTS,S alone arm, 58/162 (35.8%) in the SMC alone arm and 35/162 (21.6%) in the combined RTS,S+SMC arm.

The age, gender and country distribution of severe malaria cases are shown in Table 2. Overall, the mean age of children with severe malaria was 2.9 years (SD =1.1); 79.0% (128/162) of the cases occurred in children aged 3 years or less and the remaining 21.0% (34/162) in those 4 to 5 years.

**Table 2.** Distribution of severe malaria cases by age, gender and country.

|  | **Overall** |  | **Burkina Faso** |  | **Mali** |  |  |
| --- | --- | --- | --- | --- | --- | --- | --- |
|  | **(N=162)** |  | **(N=127)** |  | **(N=35)** |  |  |
|  | n | % | n | % | n | % | P |
| **Age (years)** |  |  |  |  |  |  | *0.002* |
| ≤1 year | 30 | 18.5 | 29 | 22.8 | 1 | 2.9 |  |
| 2 years | 62 | 38.3 | 52 | 40.9 | 10 | 28.6 |  |
| 3 years | 36 | 22.2 | 22 | 17.3 | 14 | 40.0 |  |
| 4–5 years | 34 | 21.0 | 24 | 18.9 | 10 | 28.6 |  |
| Mean (SD) | 2.93(1.11) |  | 2.80(1.07) |  | 3.42(1.12) |  | *0.003* |
| **Gender** |  |  |  |  |  |  |  |
| Male (n, %) | 103 | 63.6 | 83 | 65.4 | 20 | 57.1 | *0.37* |

The clinical presentations of severe malaria overall, and in each study arm, are presented in Table 3. Overall, the most common presentation was severe anemia reported in 81/162 children (50.0%), followed by repeated convulsions 41/162 (25.3%), prostration 18/162 (11.1%), hyperparasitaemia 13/162 (8.0%) and cerebral malaria 10/162 (6.2%). Severe malaria anemia was more frequent 62.1% (36/58) in the SMC alone arm and lowest in SMC+ RTS,S arm (37.1%, 13/35)(p = 0.048). There was no significant difference in frequencies of the other clinical presentations between the study arms (Table 3).

**Table 3.** Distribution of severe malaria presentations* by study arm.

|  | **Overall**<br>**(N=162)** |  | **RTS,S alone**<br>**(N=69)** |  | **SMC alone**<br>**(N=58)** |  | **RTS,S +SMC**<br>**(N=35)** |  |  |
| --- | --- | --- | --- | --- | --- | --- | --- | --- | --- |
|  | n | % | n | % | n | % | n | % | *P* |
| Severe malaria anemia | 81 | 50.0 | 32 | 46.4 | 36 | 62.1 | 13 | 37.1 | 0.048 |
| Repeated convulsions | 41 | 25.3 | 17 | 24.6 | 13 | 22.4 | 11 | 31.4 | 0.62 |
| Prostration | 18 | 11.1 | 10 | 14.5 | 2 | 3.4 | 6 | 17.1 | 0.063 |
| Hyperparasitaemia | 13 | 8.0 | 6 | 8.7 | 5 | 8.6 | 2 | 5.7 | 0.87 |
| Cerebral malaria | 10 | 6.2 | 5 | 7.2 | 3 | 5.2 | 2 | 5.7 | 0.92 |
| Repeated vomiting | 3 | 1.9 | 0 | 0 | 1 | 1.7 | 2 | 5.7 | 0.10 |
| Respiratory distress | 2 | 1.2 | 2 | 2.9 | 0 | 0 | 0 | 0 | 0.51 |
| Hypoglycemia | 2 | 1.2 | 1 | 1.4 | 1 | 1.7 | 0 | 0 | 0.99 |
| Jaundice | 1 | 0.6 | 0 | 0 | 1 | 1.7 | 0 | 0 | 0.57 |

*\*Note that a child can have more than one presentation*

There were marked differences in clinical phenotypes of the 162 children with severe malaria, between Burkina Faso and Mali (Table 4). Severe anemia was significantly more frequent in Burkina Faso (55.9%) compared with Mali (28.6%) (p = 0.004). Cerebral malaria also showed a marked difference, occurring in only 0.8% of cases in Burkina Faso but in 25.7% in Mali (p < 0.001). Hyperparasitaemia was absent in Burkina Faso but frequent in Mali (37.1%; p < 0.001). In contrast, convulsions and prostration did not differ significantly between countries (p = 0.41 and p = 0.12, respectively).

**Table 4.** Distribution of severe malaria presentations* by country.

|  | Overall |  | Burkina Faso |  | Mali |  |  |
| --- | --- | --- | --- | --- | --- | --- | --- |
|  | (N=162) |  | (N=127) |  | (N=35) |  |  |
|  | n | % | n | % | n | % | P |
| Severe malaria anemia | 81 | 50.0 | 71 | 55.9 | 10 | 28.6 | 0.004 |
| Repeated convulsions | 41 | 25.3 | 34 | 26.8 | 7 | 20.0 | 0.41 |
| Prostration | 18 | 11.1 | 17 | 13.4 | 1 | 2.9 | 0.125 |
| Hyperparasitaemia | 13 | 8.0 | 0 | 0.0 | 13 | 37.1 | <0.001 |
| Cerebral malaria | 10 | 6.2 | 1 | 0.8 | 9 | 25.7 | <0.001 |
| Repeated vomiting | 3 | 1.9 | 2 | 1.6 | 1 | 2.9 | 0.521 |
| Respiratory distress | 2 | 1.2 | 1 | 0.8 | 1 | 2.9 | 0.386 |
| Hypoglycemia | 2 | 1.2 | 2 | 1.6 | 0 | 0.0 | 0.99 |
| Jaundice | 1 | 0.6 | 1 | 0.8 | 0 | 0.0 | 0.99 |

\*Note that a child can have more than one presentation

The age distribution of each specific presentation of severe malaria is presented in Figure 2. Severe malaria anemia and convulsion peaked at 2 years of age while the peak for cases with prostration, hyperparasitaemia and cerebral malaria occurred at 3 years of age. The median ages of children with these clinical presentations were 2.7, 2.5, 3.3, 3.6 and 3.3 years respectively.

**Figure 2:**
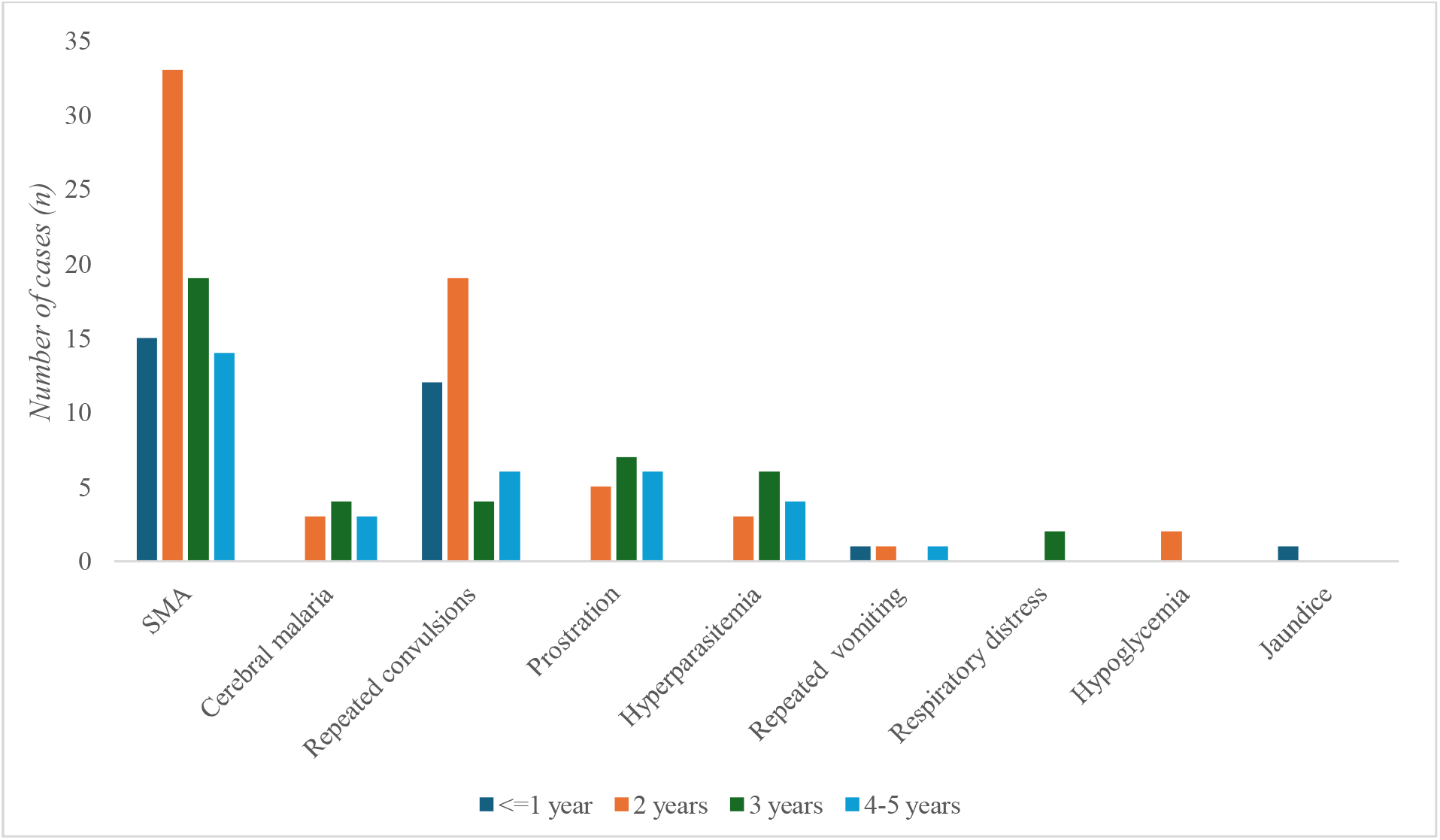
Distribution of severe malaria by age (SMA – Severe Malaria Anaemia)

The distribution of severe malaria cases by month is presented in Figure 3 showsand shows that 85.8% (139/162) of severe malaria cases occurred during the peak malaria transmission season (July to December).

**Figure 3:**
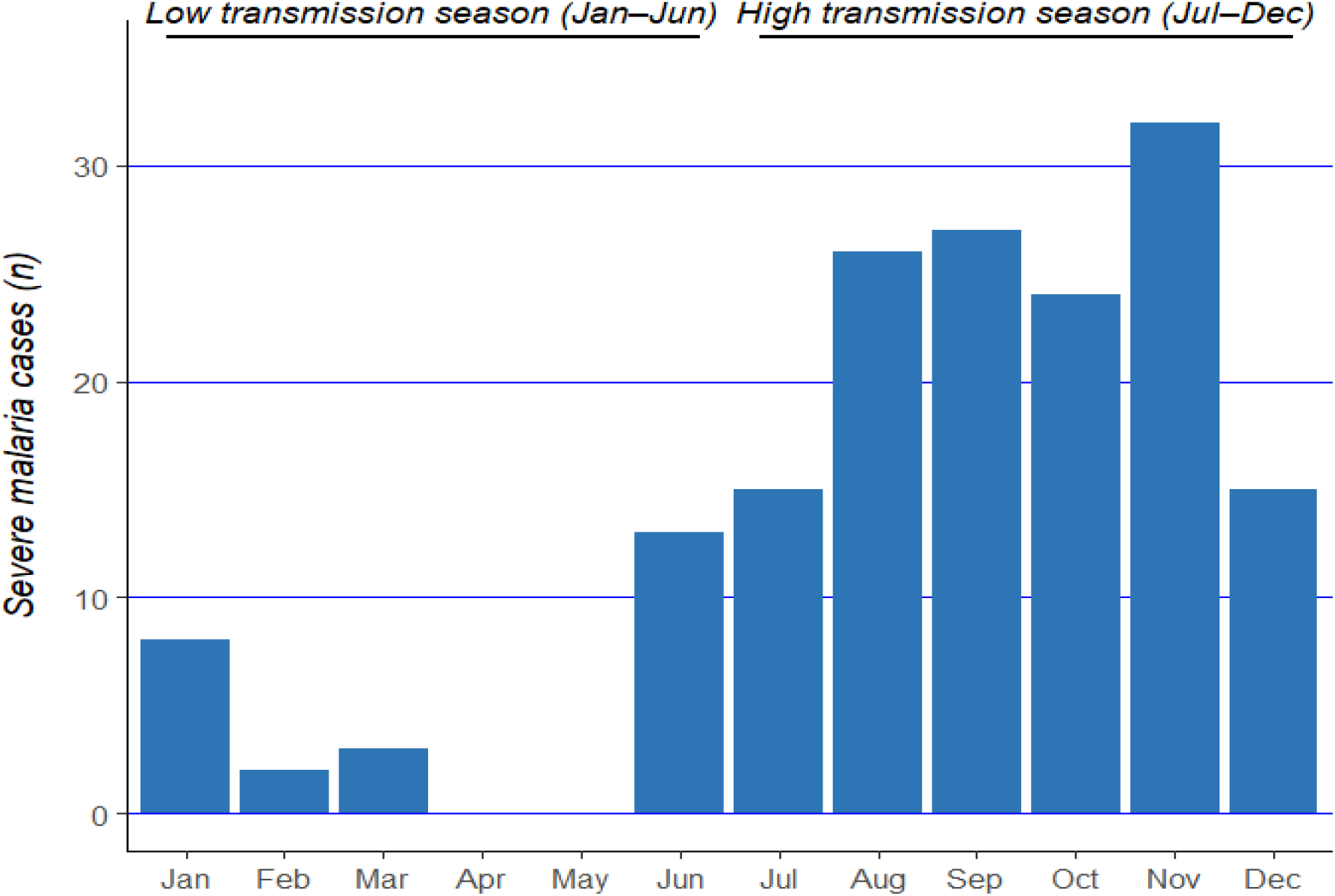
Distribution of severe malaria by month of year.

Twenty-one of the 162 children (13.0%) admitted to hospital with severe malaria cases died. The proportions were similar in Burkina Faso (15/127; 11.8%) and Mali (6/35; 17.1 %) (p= 0.40). The most frequent presentation of severe malaria in the 21 children who died was severe malaria anemia 13, followed by cerebral malaria and repeated convulsions with 4 deaths attributed to each; 1 death was due to respiratory distress and 1 to hypoglycemia.

## DISCUSSION

The clinical spectrum of severe malaria observed in this study was dominated by severe malarial anemia, accounting for nearly half of all cases, with marked differences observed across intervention arms. The highest proportion of severe malarial anemia occurred in children who received SMC alone, whereas the lowest frequency was observed in the combined RTS,S + SMC arm, with an intermediate effect in the RTS,S alone. This gradient suggests that the addition of RTS,S to SMC provided incremental protection against the development of severe anemia perhaps by decreasing the incidence of infection combined with the suppressive effect of SMC on any infections that were not prevented by the vaccine In contrast, the distribution of other severe malaria manifestations including repeated convulsions, cerebral malaria, and hyperparasitaemia did not differ significantly between arms. These features are less frequent and may be driven by acute parasite–host interactions that are less sensitive to reductions in overall infection burden. The lack of statistically significant differences for these outcomes may also reflect limited power due to relatively small numbers of cases with these clinical presentations.

Together, these findings indicate that while combined preventive strategies may not substantially alter the overall clinical pattern of severe malaria, they appear particularly effective in reducing severe malarial anemia, a key determinant of malaria-related mortality and long-term morbidity.

Severe malaria anemia was the most common clinical presentation in this study (50%), followed by repeated convulsions, prostration, hyperparasitaemia, and cerebral malaria. These findings are not consistent with previous studies conducted in different areas in Mali and Burkina Faso with higher proportions of coma and prostration ^21,22^ The age profile of children with severe malaria was consistent with existing knowledge on development of malaria immunity. Most cases occurred in those aged 3 years or less, with a mean age of 2.9 years, with specific syndromes peaking at different ages - severe anaemia at 2.8 years and cerebral malaria at 3.7 years. This pattern mirrors the immuno-epidemiological transition described in earlier studies^11,23,24^, with children progressively acquiring partial immunity, and the spectrum of clinical presentation shifting with age. A study conducted by Reyburn et al. ^10^ also found that as transmission intensity declines, the median age of severe malaria cases increases, and cerebral malaria becomes more prominent ^10^, in line with the findings of our study.

Previous studies have shown that severe malarial anemia was historically the predominant presentation of severe malaria in high-transmission settings, while cerebral malaria was more common in areas of low transmission ^10,25^. Over time, the clinical presentation of severe malaria has shifted, with a continued dominance of severe malaria anaemia, particularly in children younger than 3 years of age, due to high transmission intensity, as well as an increase in convulsions and cerebral malaria in older children, likely due to delayed immunity development as transmission declines.

The mean age of severe malaria cases in this study was 2.9 years, with differences between Mali and Burkina Faso. In Burkina Faso, the mean age was 2.8 years, while in Mali, it was 3.4 years with significant difference in proportions of severe malaria anemia, cerebral malaria and hyperpasitaemia.hyperparasitemia. These differences likely reflect in the higher malaria transmission typically resulting in earlier severe disease, and in Burkina Faso compared to Mali, and the the fact that in Mali, children had easier access to a health facility than in Burkina.

Previous studies confirm this trend in Burkina Faso where before SMC deployment, the mean age of severe malaria cases was 4.8 years in urban areas and 2.2 years in rural areas, highlighting the impact of transmission intensity on disease onset ^21^. The age-specific burden (79.0% in children aged 3 years or less) aligns with findings from Gabon (71.8% in <5 years) ^26^ but contrasts with two other studies conducted in Mozambique and Kenya ^27,28^ on the epidemiology of severe malaria, where older children increasingly contributed to severe malaria cases.

There was a seasonal concentration of cases, with over 80% of severe malaria cases occurring from July to December. This reinforces the rationale for time-aligned interventions, as implemented in the RTS,S +SMC trial and consistent with previous studies from the Sahel and sub-Sahelian regions ^12,17^. It also supports the WHO’s recommendations of aligning preventive interventions, particularly SMC and seasonal malaria vaccine delivery, with peak transmission periods to maximize impact.

Mortality among the cases of severe malaria remained high (13.0% in overall). Severe anaemia accounted for most deaths. Three of these deaths occurred after evaluation at the research health center and but before the admission in the hospital, which reflects persistent gaps in timely healthcare access; a challenge echoed in other studies ^25^. Strengthening community case detection and emergency referral systems remains essential to complement malaria prevention.

The strengths of this study include a rigorous surveillance framework, with hospital admissions recorded by trial-assigned clinical staff and severe malaria diagnoses made adhering to WHO’s standard definitions. The clinical examination and laboratory confirmation examinations by the study staff resulted in reliable reporting and classification of the presentations.

Limitations of the study include the relatively small number of cases in the combination arm, limiting the statistical power for detecting differences in syndrome distribution. Because children in this study were followed only until five years of age, cases of severe malaria occurring in older children were not captured. The study findings are specific to high seasonal transmission settings and may not be generalizable to regions with perennial or unstable transmission.

## CONCLUSIONS

Severe anaemia was the most common presentation of severe malaria in the trial population, followed by repeated convulsions. Children who received SMC alone experienced the highest proportion of severe malarial anemia, whereas the lowest frequency was observed in the combined RTS,S + SMC arm. Children aged 3 years or less were the most affected and more than four-fifths of severe malaria cases occurred between July and December, highlighting the strong seasonal concentration of severe malaria cases in the study areas.

## Data Availability

Individual de-identified data which will be held at the London School of Hygiene and Tropical Medicine (https://datacompass.lshtm.ac.uk/)and will be made available upon reasonable request.

https://datacompass.lshtm.ac.uk/

## LIST OF ABBREVIATIONS

CM: cerebral malaria
GSK: GlaxoSmithKline
ITN: insecticide-treated net
PfHRP2: *Plasmodium falciparum* histidine-rich protein 2
RTS,S: RTS,S/AS01_E_ malaria vaccine
SAE: serious adverse event
SD: standard deviation
SMA: severe malaria anaemia
SMC: seasonal malaria chemoprevention
SPAQ: sulphadoxine–pyrimethamine plus amodiaquine
WHO: World Health Organization.

## DECLARATIONS

### Ethics approval and consent to participate

Ethical approval for this study was obtained from the Ethics Committee of the University of Science, Technique, and Technology of Bamako in Mali, the Ethics Committee for Health Research in Burkina Faso and the Research Ethics Committee of the London School of Hygiene and Tropical Medicine in UK. Individual, written informed consent was obtained from the parents/guardian of study children.

### Consent for publication

Not applicable

### Authors’ contributions

The first draft of the paper was written by DI and reviewed by AD and BG. No paid support was employed in writing the paper. DC and BG coordinated the study; A Dicko, IS and JB and HT managed the field operations. DI performed the statistical analysis. IZ, YS, YDC, RSY MK, CZ, ROZ, OMD, YK, AT, ST, KS, MD, AB, AM contributed significantly to data collection; AH, IT, IK, SA and PS managed the data and reviewed the draft; PM was senior statistician of trial and reviewed the draft. JG, CKL, CFO, and OO-A contributed to the overall trial oversight and reviewed the draft. All authors read and approved the final version of the manuscript.

## Acknowledgments

We thank Karen Slater for supporting the trial in many ways; GSK for donating RTS,S/AS01_E_ and Havrix vaccines, Guilin Pharmaceuticals for supplying seasonal malaria chemoprevention drugs; the Ministry of Health and health staff in the study areas in Mali and Burkina Faso their assistance with running the trial; and all the caretakers and children for their participation.

